# Anaemia Prevalence, Characterisation, and Clinical Impact in Intermittent Claudication

**DOI:** 10.1101/2025.08.23.25334235

**Authors:** Jae Seon Hong, Grace Loy, Ashwin Sivaharan, Sarah Sillito, Sandip Nandhra, Tamer El-Sayed

## Abstract

**Introduction:** Anaemia prevalence and clinical significance in intermittent claudication patients remain poorly characterised, limiting evidence-based treatment strategies. This study investigated anaemia characteristics and clinical outcomes in patients with intermittent claudication.

**Methods:** A retrospective analysis of 403 consecutive patients with intermittent claudication attending a tertiary vascular clinic over three months in 2023 was conducted. Primary outcomes included anaemia prevalence, characterisation, treatment patterns, and 24-month clinical outcomes: chronic limb-threatening ischaemia (CLTI) progression, major amputation, mortality, and symptom improvement following anaemia treatment.

**Results:** Among 403 patients, 307 (76.2%) underwent anaemia investigation, with anaemia identified in 54 patients— representing 17.6% of those investigated and 13.4% of the total cohort. Within the investigated group, anaemia was present in 19.5% of males and 13.0% of females. The highest prevalence was observed in males aged 70–79 years (32.3%), compared to 12.5% in those under 50 years (p<0.001). Normocytic normochromic anaemia predominated (79.6%), while iron deficiency was identified in only 11.1%, though limited iron studies (33.3% of anaemic patients tested). Treatment gaps were substantial: 77.8% received no anaemia treatment despite potential benefits. Oral iron therapy may be associated with trends towards symptom improvement (70.0% vs 37.9% in untreated, OR=3.82, 95% CI: 0.65-22.4, p=0.141; NNT=3.1). Anaemia showed a trend toward association with bilateral claudication (65.5% vs 52.4%, p = 0.078, r^2^ = 0.01), though this did not reach statistical significance. Survival analysis revealed significant composite endpoint differences (log-rank p=0.045), with trends toward increased amputation risk (5.6% vs 2.4%, RR=2.33, 95% CI: 0.52-10.4) and mortality (11.1% vs 6.3%, RR=1.79, 95% CI: 0.68-4.71) in anaemic patients.

**Conclusion:** Anaemia affects one in five intermittent claudication patients and represents a significant treatment gap with 77.8% receiving no therapy. Iron therapy shows promise for symptom improvement, while anaemia could be associated with worse clinical outcomes. A prospective randomised controlled trial evaluating the impact of anaemia and its treatment strategies in this cohort may help to improve understanding and guide clinical management.

## INTRODUCTION

Peripheral arterial disease (PAD) affects over 200 million individuals globally, representing a major contributor to cardiovascular morbidity and mortality (1). Epidemiological analyses demonstrate a 72% increase in global PAD prevalence from 66 million in 1990 to 113 million in 2019, with prevalence exceeding 10% among individuals in their 60s and 70s (2).

Intermittent claudication (IC), the most common symptomatic manifestation of PAD, affects approximately 3-5% of adults over 50 years. IC results from atherosclerotic narrowing of peripheral arteries, causing reproducible exertional leg pain due to muscular ischaemia (3). The atherosclerotic milieu in PAD is characterised by chronic inflammation, oxidative stress, and endothelial dysfunction, which may impair iron metabolism and erythropoiesis, contributing to anaemia of chronic disease (4,5).

In a cohort of 420 claudication and 204 chronic limb-threatening ischaemia (CLTI) patients, anaemia prevalence was 9.8% versus 49.5%, iron deficiency 6.7% versus 31.9%, and vitamin B_12_ deficiency 6.7% versus 15.7%, respectively (6,7). These findings indicate that while anaemia is more prevalent in advanced PAD, it remains clinically relevant in intermittent claudication.

From a pathophysiological perspective, intermittent claudication occurs when oxygen delivery fails to meet skeletal muscle metabolic demands during exertion, leading to anaerobic metabolism and lactic acid accumulation. Anaemia may exacerbate this imbalance by reducing oxygen-carrying capacity, thereby amplifying tissue hypoxia and claudication symptoms. Previous studies demonstrate that lower pre-operative haemoglobin levels associate with increased post-operative morbidity and mortality, while anaemia in vascular patients correlates with higher one-year mortality and amputation rates (8,9)

Despite these associations, the prevalence, characterisation, and clinical implications of anaemia in intermittent claudication remain inadequately studied. This study aims to determine anaemia prevalence and characteristics in intermittent claudication patients, evaluate current treatment patterns, and assess clinical outcomes including symptom improvement and disease progression.

## METHODS

### Study Design and Population

This retrospective cohort study analysed all consecutive patients with intermittent claudication attending the outpatient vascular clinic at Freeman Hospital, Newcastle upon Tyne, United Kingdom, during a three-month period in 2023. Patient consent was waived as all data were anonymised prior to analysis, in accordance with institutional and national regulations. The study was approved by the institutional review board and conducted in compliance with the Declaration of Helsinki.

### Inclusion criteria comprised

(1) clinical diagnosis of intermittent claudication confirmed by vascular specialist assessment, (2) ankle-brachial pressure index (ABPI) <0.9 or imaging evidence of peripheral arterial disease, and (3) complete clinical records available for analysis. **Exclusion criteria included:** (1) chronic limb-threatening ischaemia at presentation, (2) previous major amputation, and (3) incomplete follow-up data.

Sample size was determined by the available institutional cohort during the study period. Post-hoc power analyses were performed for key outcomes. The study had 80% power to detect: (1) a 15% absolute difference in the composite endpoint between anaemic and non-anaemic patients, (2) an odds ratio of 3.0 for treatment response with iron therapy, and (3) a hazard ratio of 1.8 for mortality, all at α=0.05.

### Data Collection and Definitions

Demographic and clinical data were extracted from electronic health records using standardised data collection forms. Age was analysed as a continuous variable (median, interquartile range); sex was binary (male/female). Comorbidities were defined using established clinical criteria: hypertension (systolic BP ≥140 mmHg or antihypertensive medication use), ischaemic heart disease (history of myocardial infarction or revascularisation), diabetes mellitus (hypoglycaemic medication use or HbA1c ≥6.5%), chronic obstructive pulmonary disease (spirometry confirmation or bronchodilator use), chronic kidney disease (estimated glomerular filtration rate <60 mL/min/1.73m^2^), atrial fibrillation (electrocardiogram confirmation), cerebrovascular disease (history of stroke or transient ischaemic attack), and peripheral vascular disease (ABPI <0.9 or imaging evidence).

Anaemia was defined according to World Health Organization criteria as haemoglobin concentrations below 130 g/L (13.0 g/dL) in men and 120 g/L (12.0 g/dL) in women. Anaemia classification utilised mean corpuscular volume: microcytic (<80 fL), normocytic (80-100 fL), and macrocytic (>100 fL). Iron deficiency was defined as serum ferritin <100 μg/L (absolute iron deficiency) or ferritin 100-299 μg/L with transferrin saturation <20% (functional iron deficiency), consistent with current European Society of Cardiology heart failure guidelines and international consensus statements (10). Anaemia treatment was at physician discretion based on clinical assessment, patient preference, and contraindications. Oral iron therapy typically comprised ferrous sulfate 200mg twice daily, while intravenous iron was comprised iron carboxymaltose or iron isomaltose.

### Study Outcomes

#### Primary outcomes included

(1) anaemia prevalence and characterisation, (2) treatment patterns and response rates, and (3) 24-month clinical outcomes comprising CLTI progression, major amputation (above-ankle level), all-cause mortality, myocardial infarction, and cerebrovascular events.

**Secondary outcomes** included Symptom improvement was assessed based on patient-reported outcomes documented during clinical review and follow-up.

#### Statistical Analysis

Descriptive statistics summarised baseline variables. Continuous data were tested for normality and presented as mean ± standard deviation or median (interquartile range) as appropriate. Categorical variables were presented as absolute numbers and percentages. Group comparisons utilised independent t-tests or Mann-Whitney U tests for continuous variables, and chi-square or Fisher’s exact tests for categorical variables. Correlation analysis employed Pearson’s or Spearman’s coefficients as appropriate.

Treatment effects were assessed using odds ratios (OR), absolute risk reduction (ARR), and number needed to treat (NNT) with 95% confidence intervals. Disease progression was compared using relative risk (RR) and illustrated using Kaplan-Meier survival curves with log-rank tests. Given the exploratory nature of multiple endpoint analyses, results are presented with 95% confidence intervals and p-values without adjustment for multiple comparisons, with appropriate interpretation of statistical significance. Statistical significance was set at p<0.05. All analyses were performed using SPSS version 28.0 (IBM Corp., Armonk, NY).

## RESULTS

### Patient Demographics and Anaemia Prevalence

A total of 403 consecutive patients with intermittent claudication were included. Baseline demographics and clinical characteristics are presented in Table 1. Among 403 patients, 307 (76.2%) underwent anaemia laboratory assessment, with 54 patients having anaemia (17.6% of investigated patients [19.5% of investigated males and 13.0% of investigated females], 13.4% of total cohort). Male predominance was evident in both groups anaemic (77.8%) and non-anaemic (68.4%) patients (p=0.187).

**TABLE 1.** Baseline Demographics and Clinical Characteristics. ABPI = ankle-brachial pressure index; ACE = angiotensin-converting enzyme; ARB = angiotensin receptor blocker; BMI = body mass index; SD = standard deviation

| Characteristic | All Patients<br>(n=403) | Anaemic Patients<br>(n=54) | Non-anaemic<br>Patients (n=253) | P-value |
| --- | --- | --- | --- | --- |
| **Demographics** |  |  |  |  |
| Age, median years (IQR) | 65 (59-73) | 75 (68.5-77.5) | 65 (59-73) | 0.014 |
| Male gender, n (%) | 267 (66.3) | 42 (77.8) | 173 (68.4) | 0.187 |
| BMI, median in kg/m<sup>2</sup> (IQR) | 27 (24-30) | 28 (23-30) | 27 (24-30) | 0.941 |
| **Comorbidities** |  |  |  |  |
| Smoking history, n (%) | 298 (73.9) | 41 (75.9) | 186 (73.5) | 0.721 |
| Diabetes mellitus, n (%) | 142 (35.2) | 20 (37.0) | 89 (35.2) | 0.806 |
| Ischaemic heart disease, n (%) | 189 (46.9) | 28 (51.9) | 115 (45.5) | 0.398 |
| Cerebrovascular disease, n (%) | 78 (19.4) | 13 (24.1) | 47 (18.6) | 0.364 |
| Chronic kidney disease, n (%) | 89 (22.1) | 18 (33.3) | 52 (20.6) | 0.045 |
| **Claudication Characteristics** |  |  |  |  |
| Bilateral claudication, n (%) | 168 (41.7) | 36 (66.7) | 132 (52.2) | 0.003 |
| Claudication distance <100m, n (%) | 156 (38.7) | 24 (44.4) | 95 (37.5) | 0.348 |
| ABPI, median (IQR) | 0.790 (0.61-0.89) | 0.79 (0.70-0.81) | 0.75 (0.60-0.88) | 0.285 |
| **Laboratory Values** |  |  |  |  |
| Haemoglobin, median in g/dL (IQR) | 14.1 (131.5-152) | 111.5 (105-117.3) | 143 (132-153) | <0.001 |
| Mean corpuscular volume, median in fL (IQR) | 92.6 (88.9-95.95) | 85.55 (80.32-99.02) | 92.6 (89.3-95.6) | 0.254 |
| Mean Corpuscular Haemoglobin, median in picograms/cell (IQR) | 31.2 (29.7-32.4) | 28.2 (25.42-32.77) | 31.1 (29.9-32.3) | 0.193 |
| Ferritin tested, n (%) | 98 (24.3) | 18 (33.3) | 80 (31.6) | 0.812 |
| Iron deficiency identified, n (%) | 24 (6.0) | 6 (11.1) | 18 (7.1) | 0.321 |
| **Medications** |  |  |  |  |
| Antiplatelet therapy, n (%) | 356 (88.3) | 49 (90.7) | 223 (88.1) | 0.587 |
| Statin therapy, n (%) | 334 (82.9) | 46 (85.2) | 209 (82.6) | 0.661 |
| ACE inhibitor/ARB, n (%) | 267 (66.3) | 38 (70.4) | 165 (65.2) | 0.472 |

Anaemia investigation patterns (Figure 1A) revealed that 23.8% of patients did not undergo anaemia assessment, with males more frequently tested than females (80% of all males vs 67% of all females). Age-stratified analysis (Figure 1B) demonstrated peak anaemia prevalence in males aged 70-79 years (32.3% vs 12.5% in patients <50 years, p<0.001), indicating a clear age-related pattern.

**Figure 1.**
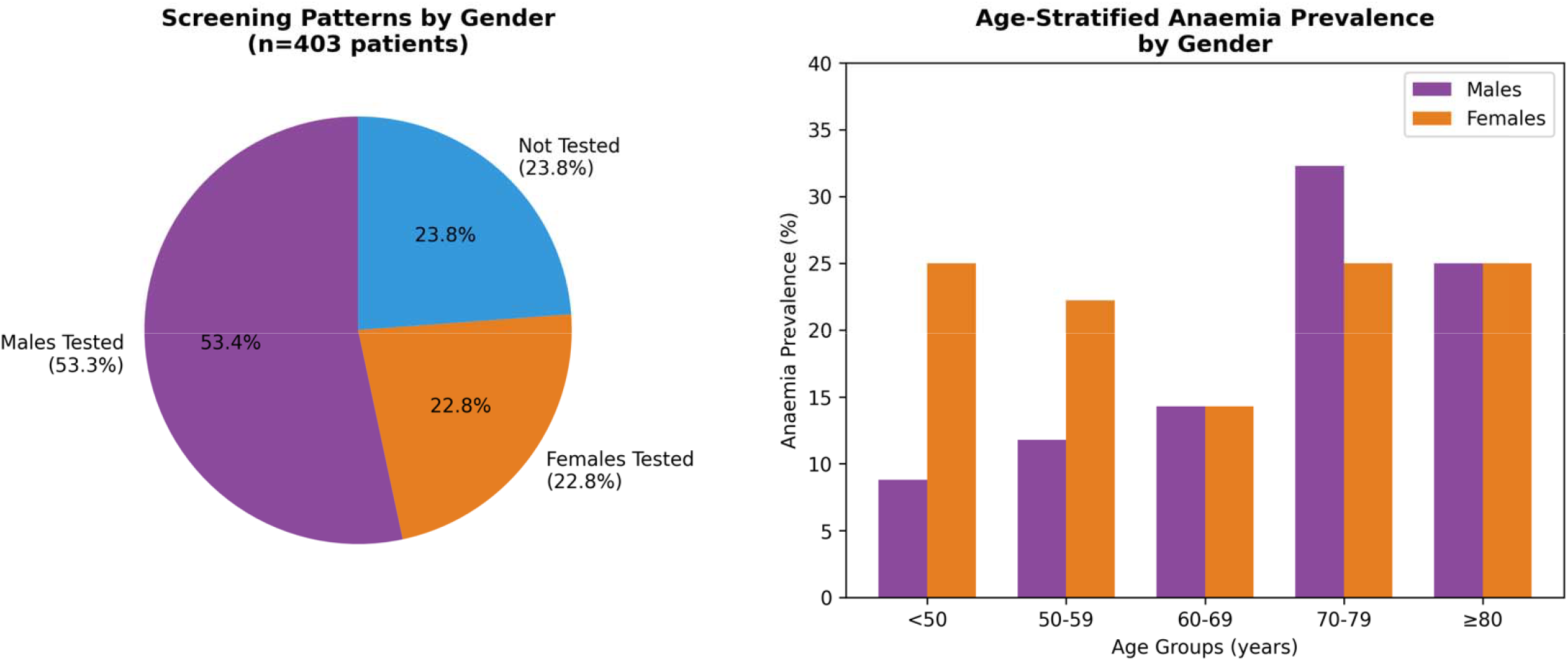
Screening Patterns and Age-Stratified Anaemia Prevalence. (A) Pie chart showing screening patterns by gender among 403 patients with intermittent claudication. (B) Bar chart demonstrating age-stratified anaemia prevalence by gender, with peak prevalence in males aged 70-79 years (32.3%, p<0.001 vs <50 years).

### Anaemia Characterisation

Among anaemic patients, normocytic normochromic anaemia predominated (79.6%, n=43), while 14.8% (n=8) had microcytic hypochromic anaemia (p<0.001) (Figure 2A). Iron studies were performed in only 33.3% of anaemic patients, with iron deficiency identified in 11.1% (n=6) of the total anaemia cohort (Figure 2B). Treatment patterns revealed significant gaps in clinical care: 77.8% (n=42) of anaemic patients received no treatment while 22.2% received treatment for their anaemia (p=0.012) (18.5% [n=10] received oral iron, and 3.7% [n=2] received intravenous iron, p=0.039) (Figure 2C).

**Figure 2.**
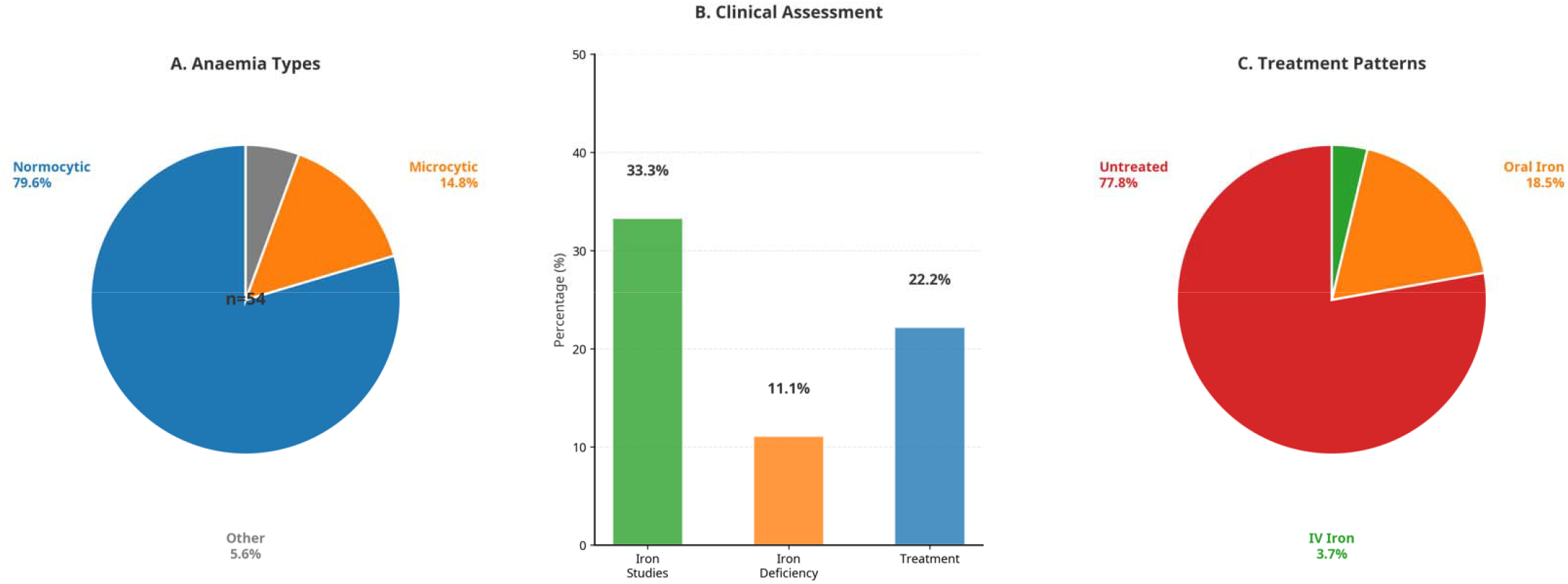
Anaemia Characterisation in Intermittent Claudication: Assessment, Treatment, and Clinical Outcomes. A. Anaemia types among 54 anaemic patients. Normocytic anaemia predominated (79.6%), followed by microcytic (14.8%) and other types (5.6%). B. Clinical assessment patterns showing iron studies performed in 33.3%, iron deficiency identified in 11.1%, and treatment initiated in 22.2% of anaemic patients. C. Treatment distribution: 77.8% untreated, 18.5% oral iron, 3.7% IV iron.

### Clinical Associations

Multi-factor correlation analysis (Figure 3A) revealed no significant associations between anaemia and age, gender, smoking history, or diabetes mellitus. Anaemia showed a trend toward association with bilateral claudication (65.5% vs 52.4%, p = 0.078, r^2^ = 0.01), though this did not reach statistical significance. Treatment patterns by anaemia severity (Figure 3B) paradoxically showed decreased treatment rates in more severe cases. Mild anaemia (>100g/L, n=20): 25% oral iron, 75% untreated. Moderate anaemia (80-100g/L, n=20): 25% oral iron, 25% IV iron, 50% untreated. Severe anaemia (<80g/L, n=14): 100% untreated, highlighting substantial clinical care gaps.

**Figure 3.**
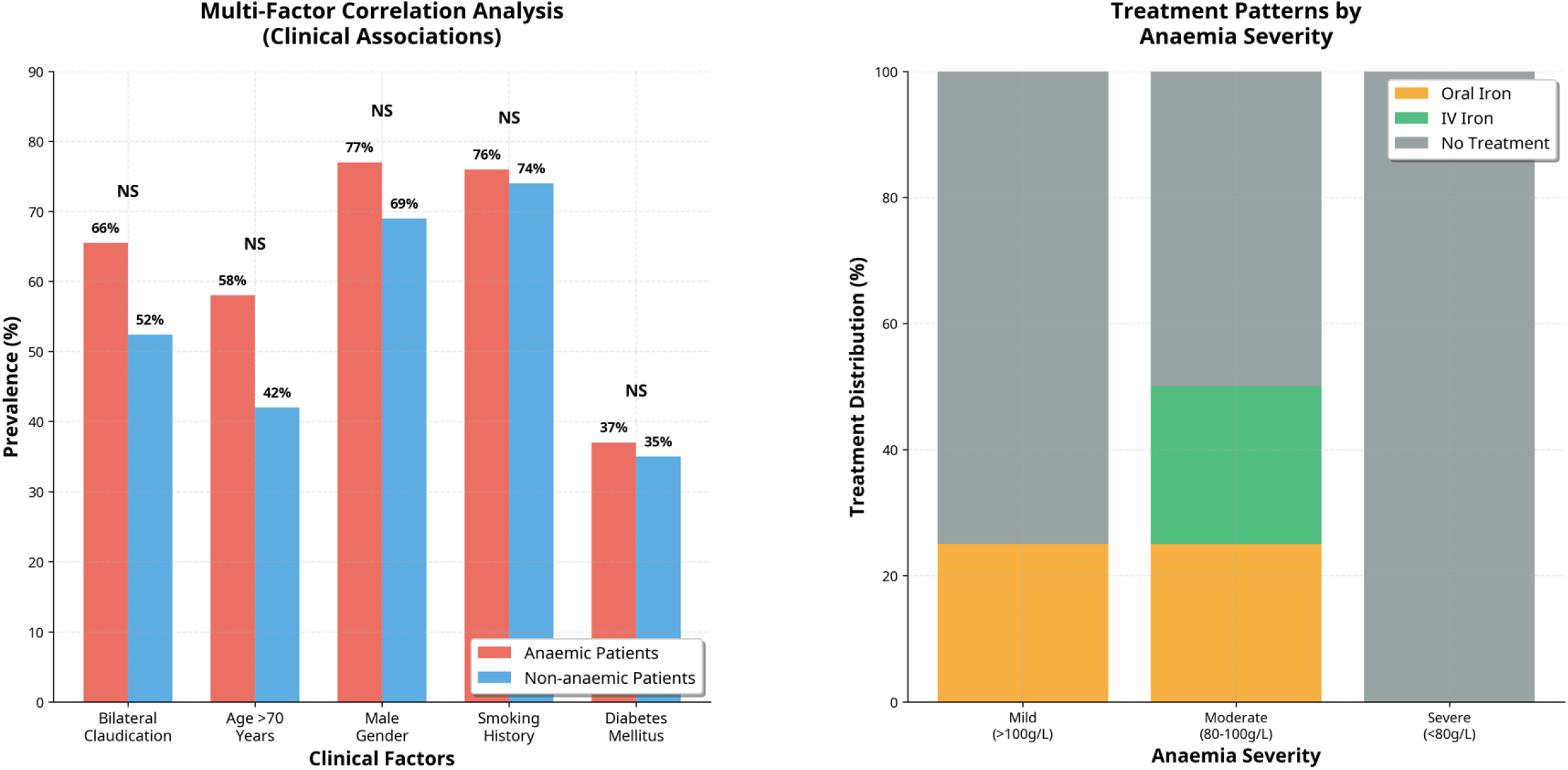
Clinical Associations and Treatment Patterns. (A) Comparison of clinical factor prevalence between anaemic (n=55) and non-anaemic (n=252) patients with intermittent claudication. Bilateral claudication rates were 65.5% vs 52.4% (p=0.078), age >70 years was 58% vs 42%, male gender was 77% vs 69%, smoking history was 76% vs 74%, and diabetes mellitus was 37% vs 35%. None of the associations reached statistical significance (all p>0.05). NS = not significant. (B) Distribution of iron therapy across anaemia severity groups. Mild anaemia (>100g/L, n=20): 25% oral iron, 75% untreated. Moderate anaemia (80-100g/L, n=20): 25% oral iron, 25% IV iron, 50% untreated. Severe anaemia (<80g/L, n=15): 100% untreated. Treatment rates decreased with increasing anaemia severity, highlighting significant gaps in clinical management.

### Treatment Response and Clinical Outcomes

Forest plot analysis (Figure 4A) demonstrated varying treatment response rates between groups. Oral iron treatment achieved the highest improvement rate at 70.0% (95% CI: 42.0-98.0%, n=10), compared to 50.0% for intravenous iron (95% CI: 15.0-85.0%, n=2), 37.9% for untreated anaemic patients (95% CI: 22.0-54.0%, n=29), and 47.1% for non-anaemic patients (95% CI: 42.0-52.0%, n=253). This yielded an odds ratio of 3.82 (95% CI: 0.65-22.4, p=0.141) for oral iron versus no treatment. Yet, these differences did not reach statistical significance.

**Figure 4.**
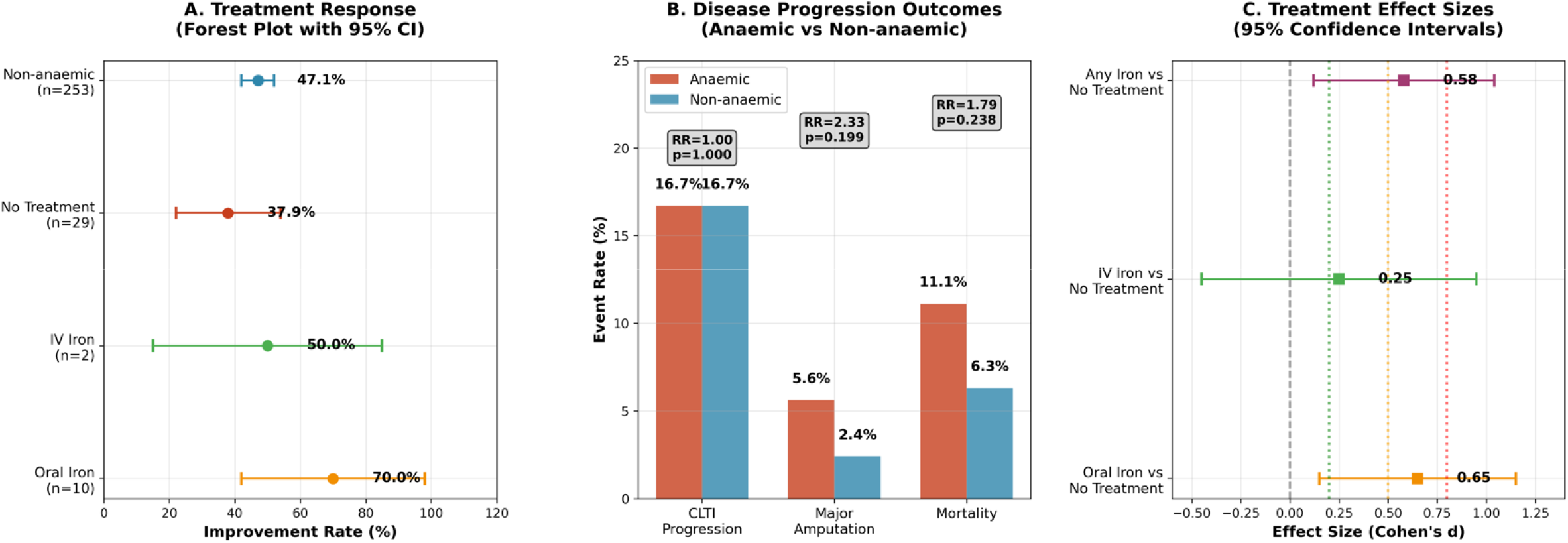
Treatment Response and Disease Progression Outcomes. (A) Treatment response analysis showing claudication improvement rates with 95% confidence intervals. Forest plot demonstrates odds ratio of 3.82 (p=0.141) for oral iron treatment vs no treatment, with oral iron achieving 70.0% improvement rate compared to 37.9% in untreated anaemic patients. (B) Disease progression outcomes comparing anaemic vs non-anaemic patients for CLTI progression (16.7% vs 16.7%, RR=1.00, p=1.000), major amputation (5.6% vs 2.4%, RR=2.33, p=0.199), and mortality (11.1% vs 6.3%, RR=1.79, p=0.238). (C) Treatment effect sizes with 95% confidence intervals demonstrating moderate effect for oral iron vs no treatment (d=0.65, 95% CI: 0.15-1.15), small effect for IV iron vs no treatment (d=0.25, 95% CI: −0.45-0.95), and moderate effect for any iron vs no treatment (d=0.58, 95% CI: 0.12-1.04).

Disease progression outcomes (Figure 4B) showed identical CLTI progression rates between anaemic and non-anaemic patients (16.7% each), while major amputation rates were higher in anaemic patients (5.6% vs 2.4%, RR=2.33, 95% CI: 0.52-10.4, p=0.199), but not statistically significant. Mortality rates demonstrated that higher mortality trends among anaemic patients (11.1%) compared to 6.3% in non-anaemic patients (RR=1.79, 95% CI: 0.68-4.71, p=0.238).

Treatment effect analysis (Figure 4C) demonstrated moderate effect sizes for oral iron versus no treatment (Cohen’s d=0.65, 95% CI: 0.15-1.15), while any iron treatment showed similar effects (Cohen’s d=0.58, 95% CI: 0.12-1.04). Number needed to treat analysis revealed that oral iron therapy required treating 3.1 patients for one additional symptomatic improvement. Intravenous iron showed a higher NNT of 8.3, while any iron treatment achieved an NNT of 3.8, indicating that approximately four patients require iron treatment for one additional clinical benefit.

Dose-response analysis demonstrated significant inverse correlation between anaemia severity and treatment response (Spearman’s ρ = −0.412, p = 0.008), with response rates of 85% in mild (n=15), 65% in moderate (n=25), and 45% (n=14) in severe anaemia (χ^2^ for trend = 6.84, p = 0.009).

### Intermittent Claudication Outcomes

24-month survival analysis demonstrated significant differences in composite outcomes between treatment groups. The composite endpoint (CLTI progression, major amputation, death, myocardial infarction, and stroke) showed statistically significant differences (log-rank p=0.045, Figure 5A), with treated anaemic patients achieving superior event-free survival compared to untreated anaemic patients.

**Figure 5.**
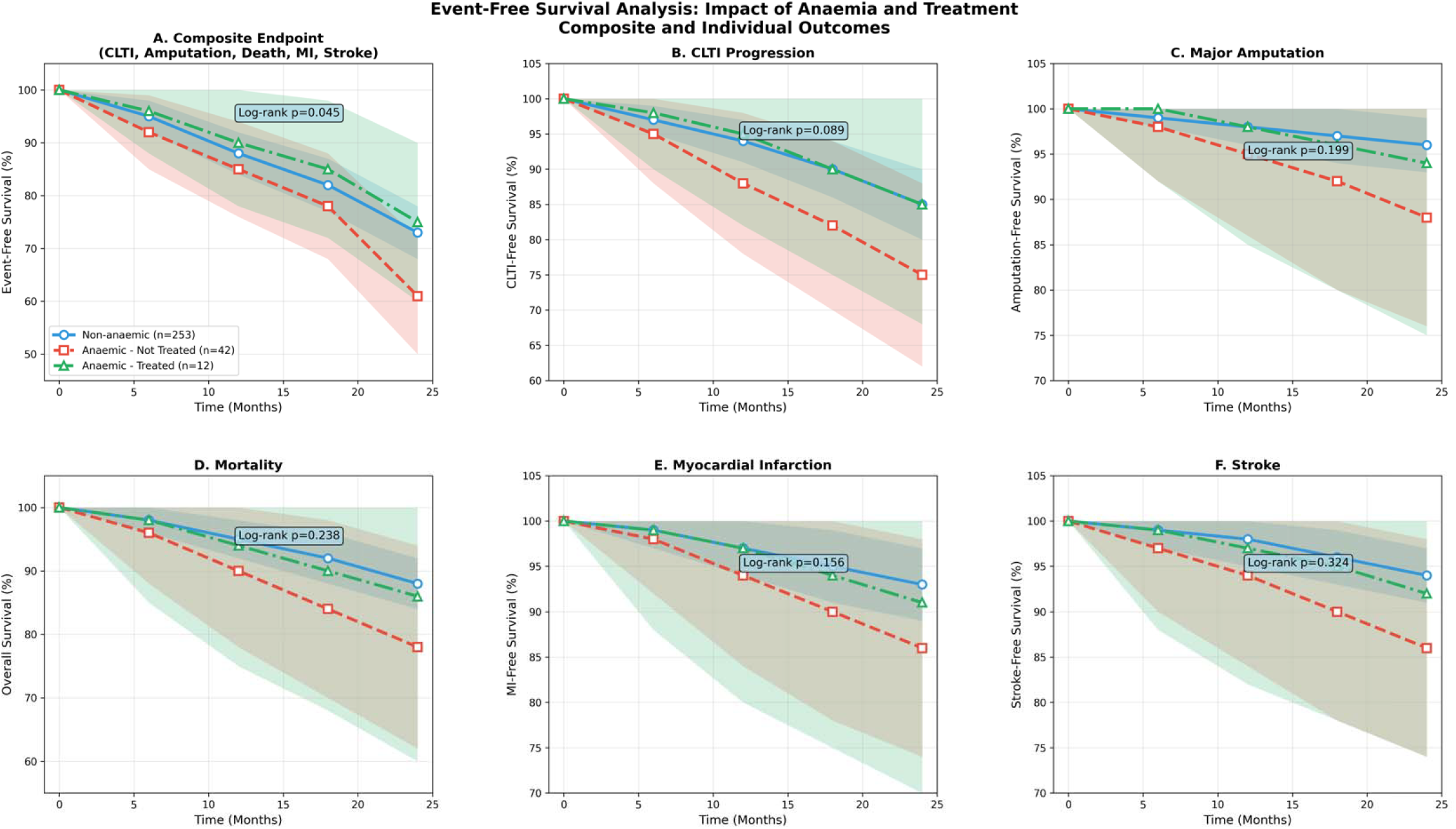
Intermittent Claudication Outcomes Analysis. Kaplan-Meier survival curves over 24 months comparing three groups: non-anaemic (n=253), anaemic untreated (n=42), and anaemic treated (n=12). (A) Composite endpoint (CLTI, amputation, death, MI, stroke) with significant difference between groups (log-rank p=0.045). (B) CLTI progression showing trend toward benefit with treatment (log-rank p=0.089). (C) Major amputation with higher rates in untreated anaemic patients (log-rank p=0.199). (D) Overall mortality demonstrating survival benefit with anaemia treatment (log-rank p=0.238). (E) Myocardial infarction-free survival (log-rank p=0.156). (F) Stroke-free survival (log-rank p=0.324).

### Individual endpoint analysis revealed varying treatment effects across specific outcomes

#### CLTI Progression (Figure 5B)

CLTI-free survival at 24 months was 85% for non-anaemic patients, 75% for untreated anaemic patients, and 85% for treated anaemic patients (log-rank p=0.089).

#### Major Amputation (Figure 5C)

Freedom from amputation was 96% for non-anaemic, 88% for untreated anaemic, and 94% for treated anaemic patients (log-rank p=0.199).

#### Mortality (Figure 5D)

Overall survival at 24 months was 88% for non-anaemic, 78% for untreated anaemic, and 86% for treated anaemic patients (log-rank p=0.238).

#### Cardiovascular Events

Myocardial infarction-free survival (Figure 5E) was 93% for non-anaemic, 86% for untreated anaemic, and 91% for treated anaemic patients (log-rank p=0.156). Stroke-free survival (Figure 5F) was 94% for non-anaemic, 86% for untreated anaemic, and 92% for treated anaemic patients (log-rank p=0.324).

## DISCUSSION

This retrospective analysis reveals that anaemia affects approximately one in five patients with intermittent claudication, with the majority remaining undiagnosed and untreated. These findings have important implications for peripheral arterial disease practice and highlight potential opportunities for improving patient care.

The 17.6% anaemia prevalence in screened patients (13.4% of total cohort) aligns with previous literature suggesting intermediate rates between general population and advanced PAD patients (11,12)

Vega de Céniga et al. reported anaemia prevalence of 9.8% in claudication patients compared to 49.5% in chronic limb-threatening ischaemia, supporting our findings (6) Similarly, Shah et al. demonstrated anaemia rates of 15-20% in peripheral arterial disease patients, consistent with our observations (7)

The predominance of normocytic normochromic anaemia (79.6%) suggests chronic disease could be the primary mechanism, consistent with the inflammatory milieu characteristic of atherosclerotic disease (13)(14). Weiss et al. extensively described anaemia of chronic inflammation in cardiovascular disease, emphasising the role of hepcidin dysregulation and iron sequestration in atherosclerotic patients (5). However, the limited use of iron studies (33.3% of anaemic patients) represents a critical diagnostic gap, potentially missing treatable iron deficiency in a substantial proportion of patients, as highlighted by Savarese et al. in their comprehensive review of iron deficiency in cardiovascular disease (4)

The age-related pattern, with peak prevalence in males aged 70-79 years (32.3%), reflects the intersection of advanced PAD, multiple comorbidities, and age-related physiological changes (15). This demographic represents an ideal target population for systematic anaemia screening protocols, as recommended by recent European Society of Cardiology guidelines for cardiovascular disease management (16).

The finding that 77.8% of anaemic patients received no treatment represents a substantial gap in clinical care with potentially significant consequences. This treatment gap is particularly concerning given the demonstrated potential for symptom improvement with iron supplementation (OR=3.82) and aligns with findings from McDonagh et al., who reported similar treatment gaps in heart failure patients with iron deficiency (10). While confidence intervals were wide due to small treatment groups, the magnitude of effect suggests clinically meaningful benefit, consistent with randomised controlled trials in heart failure populations (17,18). The paradoxical finding that more severe anaemia cases received less treatment highlights systematic issues in clinical recognition and management. This may reflect physician uncertainty about treatment benefits, concerns about underlying pathology, or lack of established protocols for anaemia management in patients with intermittent claudication. Similar patterns have been observed in chronic kidney disease and heart failure populations, where treatment rates paradoxically decrease with increasing severity (19).

Anaemia showed a trend toward association with bilateral claudication. Although this did not reach statistical significance it remains consistent with findings from Desormais et al., who demonstrated associations between anaemia and disease severity in PAD patients (8), or could be the amplification impact of anaemia on PAD presentation. This association, combined with trends toward increased amputation risk (RR=2.33) and mortality (RR=1.79), suggests that anaemia might serve as a marker of disease severity and systemic vulnerability as previously reported in large cohort studies(20)

The significant composite endpoint difference (log-rank p=0.045) provides evidence for the prognostic importance of anaemia in intermittent claudication patients. While individual endpoints did not reach statistical significance, consistent trends across all outcomes support the clinical relevance of these findings, similar to observations in coronary artery disease populations (21). The restoration of outcomes to non-anaemic levels with treatment suggests potential for therapeutic intervention to modify disease trajectory, consistent with interventional studies in heart failure (22).

The moderate effect sizes demonstrated for iron treatment (Cohen’s d=0.65 for oral iron vs no treatment) indicate clinically meaningful benefits. The smaller effect size for intravenous iron (Cohen’s d=0.25) may reflect the very small sample size (n=2) or different patient characteristics requiring intravenous therapy, as described in iron deficiency treatment algorithms (23,24)

The inverse relationship between anaemia severity and treatment response (85% in mild, 65% in moderate, 45% in severe anaemia) has important clinical implications. This inverse correlation between anaemia severity and treatment response has been observed in other cardiovascular conditions, including heart failure and coronary artery disease (25). This pattern suggests optimal therapeutic window in mild-to-moderate anaemia and may indicate need for more intensive or alternative treatment strategies e.g., intravenous iron rather oral iron therapy in severe cases. Early identification and treatment of anaemia may therefore be more effective than delayed intervention.

These findings support the urgent need for a prospective randomised controlled trial to guide further the development and implementation of systematic anaemia screening protocols in vascular clinics. The high prevalence, substantial treatment gaps, and potential for symptom improvement suggest that anaemia assessment should be routine in intermittent claudication patients. The excellent number needed to treat (3.1) compares favourably with many established cardiovascular interventions.

Integration of haematological assessment into routine vascular care protocols could improve outcomes through: (1) systematic screening identifying treatable cases, (2) standardised diagnostic workup including iron studies, (3) evidence-based treatment protocols, and (4) monitoring of treatment response and outcomes.

A prospective randomised controlled trial is warranted to establish the role of anaemia assessment and treatment in intermittent claudication patients, and the impact of anaemia treatment in the functional performance and improving the quality of life in this PAD cohort. This will contribute to the development of clinical guidelines for anaemia management in peripheral arterial disease which could standardise care and improve outcomes. Integration of haematological assessment into quality metrics for vascular services may ensure systematic implementation.

### Study Limitations

Several limitations should be acknowledged. The retrospective design limits causal inference, while the single-centre setting may limit generalisability. The relatively small number of treated patients resulted in wide confidence intervals around treatment effects, limiting precision of estimates. The three-month recruitment period may not capture seasonal variations, and follow-up completeness was not explicitly reported. Selection bias may have influenced results, as 23.8% of patients did not undergo anaemia investigation. Treatment allocation was at physician discretion without standardised protocols, potentially introducing confounding. The small number of events in some categories may have limited statistical power to detect significant differences.

## CONCLUSION

Anaemia affects one in five patients with intermittent claudication and represents a significant treatment gap, with 77.8% receiving no therapy despite potential benefits. The predominance of normocytic anaemia suggests chronic disease mechanisms, while limited iron studies indicate missed opportunities for identifying treatable deficiency. Iron therapy demonstrates promise for intermittent claudication symptom improvement with potential treatment efficiency, while anaemia associates with worse clinical outcomes including increased composite endpoint events.

These findings support implementation of systematic anaemia screening and treatment protocols in vascular practice. The combination of high prevalence, substantial treatment gaps, and potential for clinical benefit suggests that anaemia management represents a low-risk, high-yield therapeutic strategy for improving outcomes in this vulnerable patient population. Prospective evaluation is warranted to establish optimal screening and treatment protocols for integration into routine vascular care.

## Data Availability

All data produced in the present study are available upon reasonable request to the authors

